# Increased Donor Inhibitory KIR Are Associated with Reduced GVHD and Improved Survival Following HLA Matched Unrelated Donor HCT in Pediatric Acute Leukemia

**DOI:** 10.1101/2023.12.14.23299955

**Authors:** Elizabeth Krieger, Rehan Qayyum, Amir Toor

**Author notes:** Corresponding author: Amir A. Toor MD, Topper Cancer Institute, Lehigh Valley Health Network 1240 Cedar Crest Blvd., Allentown, PA 18013. Corresponding author: Elizabeth Krieger, MD. Children’s Hospital of Richmond at VCU 1000 East Broad Street, Richmond, Virginia 23219.

## Abstract

Killer immunoglobulin-like receptor (KIR) and KIR-ligand (KIRL) interactions play an important role in natural killer cell-mediated effects after hematopoietic stem cell transplantation (HCT). Previous work has shown that accounting for known KIR-KIRL interactions may identify donors with optimal NK cell-mediated alloreactivity in the adult transplant setting.

Pediatric acute leukemia patients were retrospectively analyzed and KIR-KIRL combinations and maximal inhibitory KIR ligand (IM-KIR) scores determined. Clinical outcomes were examined using a series of graphs depicting clinical events and endpoints.

The graph methodology demonstrated that prognostic variables significant in the occurrence of specific clinical endpoints, remained significant for relevant downstream events. KIR-KIRL combinations were significantly predictive for reduced grade 3-4 aGVHD likelihood, in patients transplanted with increased inhibitory KIR gene content and IM-KIR=5 scores. Improvements were also observed in associated outcomes for both ALL and AML patients, including relapse-free survival, GRFS, and overall survival.

This study demonstrates that NK cell KIR HLA interactions may be relevant to the pediatric acute leukemia transplant setting. Reduction in aGVHD suggests KIR effects may extend beyond NK cells. Moving forward clinical trials utilizing donors with a higher iKIR should be considered for URD HCT in pediatric recipients with acute leukemia to optimize clinical outcomes.

## Introduction

Clinical outcomes following allogeneic hematopoietic stem cell transplantation (HCT) are varied, depending on donor and recipient characteristics. Further complexity is introduced into analytic schemas by the different paths to survival an individual may take, e.g., with chronic GVHD or without it, after relapse or significant infection. Natural killer (NK) cells play a critical role in many of the above complications of HCT, such as, contributing to relapse and infection prevention given their early reconstitution following HCT. NK cells fulfill this important immune reconstitution function in part because they possess an array of activating and inhibitory receptors on their surface, the interactions of which may impact relapse and survival after HCT. These, killer immunoglobulin-like receptors (KIR) target non-antigen-binding domains of HLA class I molecules, KIR-ligands (KIRL). NK cell mediated alloreactivity, exemplified by KIR-KIRL interactions enables early graft versus leukemia (GVL) effects in a HLA-matched environment before T cell-mediated GVL effects might emerge. Previous studies have attempted to identify the optimal donor selection method based on donor KIR in recipients who have acute myelogenous leukemia (AML) or acute lymphoblastic leukemia (ALL), but inconsistent findings have prevented a consensus from being reached^1,2^.

In vitro inhibitory KIR when engaged have suppressive effect on NK cell function, with direct proportionality between the number of KIR engaged by KIRL and the inhibition of NK cell mediated target cell killing ^3^. Conversely activating KIR-KIRL interaction promotes granzyme and perforin release and target cell destruction in in vitro assays^4^. These interactions are compounded by an unusual property of NK cells, termed licensing, where an overall inhibitory environment is necessary for optimal killing to occur through a variety of different interactions, and the average state of an NK cell is defined by the sum of these inhibitory and activating signals^5^. As such in the absence of inhibitory KIR-KIRL interactions the killing effect of activating receptors is diminished, akin to the phenomenon of exhaustion in T cells ^6^.

In line with above logic, clinical studies in allograft recipients have focused on this question from the frame of reference of activating KIR molecules. The dominant narrative to emerge from these studies is that of the KIR A and B haplotypes, with the latter being more variable and richly endowed with activating KIR. The KIR locus on chromosome 19 is a highly polymorphic locus, with variability only exceeded by the major histocompatibility locus on chromosome 6. Studies examining KIR genotype effect on transplant led to the adoption of KIR genotyping as an additional tool for unrelated hematopoietic cell donor selection, however, because of inconsistent clinical results KIR genotyping has yet to become established as a criterion for unrelated donor selection. However, if the KIR and KIRL genotype is examined in a strictly quantitative manner to quantify KIR-KIRL interactions one may achieve different results than looking at each KIR gene individually or at the KIR haplotype in aggregate. This was done in a cohort of adult patients for whom these data were available, using a system of matrix, vector-operator equations allow for the accounting of known donor-recipient KIR-KIRL (HLA) interactions. This analysis revealed a hitherto fore unrecognized effect of donor inhibitory KIR content, such that Haplotype B/X donors with the greatest number of inhibitory KIR genes demonstrated a decrease in relapse risk following HLA-matched unrelated donor (MUD) HCT, both in the presence of their HLA ligands (iKIR) or when these ligands were missing (mKIR)^7^. This approach was then validated in a CIBMTR analysis of an adult cohort with AML^8^. The current study will explore whether this new mathematical approach to quantify KIR-KIRL combinations will be predictive of clinical outcomes in a cohort of pediatric patients who underwent HLA-MUD, T cell replete transplants. In order to gauge the relative impact of the KIR effect on outcomes following transplantation a new graph theory based analysis will be performed.

## Methods

### KIR-KIRL interaction scores

The study population consisted of pediatric donor-recipient pairs (DRP) transplanted for acute leukemia between 2005 and 2016 and were part of the Center for International Blood and Marrow Transplant Research registry. This data set was made publicly available for secondary analyses by the CIBMTR following publication of the initial findings^9^. All donors were unrelated, and transplants were facilitated via the National Marrow Donor Program. Human leukocyte antigen (HLA) match grade was 8/8, including HLA-A, -B, -C and -DRB1, with HLA typing performed at high resolution. KIR/HLA ligand status (A3, A11, Bw4 or Bw6, C1 or C2) was determined as previously reported^10,11^. KIR genotyping was available for all donors and was performed as previously described^12^ KIR-KIRL interaction scores were assigned as previously described^7,8^. In brief, KIR-KIRL interaction scores, reflecting genotypically determined, known possible KIR-HLA interactions were assigned as previously described. KIR with established HLA/KIRL were utilized^7^. An inhibitory KIR (iKIR) interaction is assigned when the donor possesses an inhibitory KIR and the recipient its known inhibitory KIRL for each DRP; iKIR were summed to generate the iKIR score. Conversely, the sum of the iKIR present in the donor without the cognate ligands in the recipient constituted the missing KIR score (mKIR). Activating KIR in the donor with their known ligands constituted the aKIR score. Activating KIR in the donor without their known ligand in the recipient were not considered in this system as there is no consensus on off HLA targets and absence of an activating KIR ligand will not affect the activation state of the NK cell. The iKIR and mKIR scores, which are mutually exclusive for a given donor, were summed to calculate the total, inhibitory donor KIR content, or inhibitory ‘maxima’, IM-KIR score for that donor; notably, IM-KIR score is equivalent to the number of donor’s inhibitory KIR genes which are known to interact with HLA.

### Statistical methods

Survival, relapse, relapse-free survival (RFS), transplant-related mortality, acute graft vs host disease (aGVHD) and chronic GVHD (cGVHD) were defined per CIBMTR criteria^13^. Relapse free & chronic GVHD free survival (GRFS) was also evaluated. To evaluate the influence of confounding variables on the effect of NK cell-mediated alloreactivity as determined by the KIR component scores, a multivariate Cox proportional hazards model was constructed including available clinically relevant variables (**Table 1**). To allow for competing events, competing risk analyses were conducted using the Fine and Gray methods. Competing risk analysis treated TRM as a competing risk for relapse and *vice versa*; for cGVHD occurrence, relapse and TRM before cGVHD diagnosis were treated analogously.

**Table 1.** Study population demographics.

| **VARIABLE** | **Study population§**<br>**(N=696)** |
| --- | --- |
| **HLA 8/8 Match** | 696 (100) |
| **AML** | 336 (48) |
| **ALL** | 360 (52) |
| **Disease Status** |  |
| CR1 | 301 (43) |
| CR2 | 254 (36) |
| CR3+ | 68 (10) |
| Active | 69 (10) |
| Missing | 4 (0.57) |
| **Recipient Age** |  |
| 0-9 | 295 (42) |
| 10-19 | 401 (58) |
| **KPS 90-100** | 570 (83) |
| **Conditioning Intensity** |  |
| Myeloablative | 632 (91) |
| RIC | 17 (2) |
| NMA | 47 (7) |
| **GVHD Prophylaxis** |  |
| Tacrolimus-Based | 376 (54) |
| Cyclosporine-Based | 320 (46) |
| **In Vivo T-Cell Depletion** | 178 (26) |
| **Ethnicity** |  |
| Non-white | 238 (34) |
| white | 458 (66) |
| **Graft Source** |  |
| Bone Marrow | 537 (77) |
| PBMC | 159 (23) |
| **IMScore=5** | 265 (38) |
| **Relapse** | 193 (28) |
| **Death** | 278 (40) |

### Clinical outcome graphs

To examine the relative impact of variables influencing clinical outcomes, graphs depicting clinical outcomes were utilized. Graphs are mathematical constructs in which nodes, or vertices are connected with edges which may be unidirectional or bidirectional. This form of network analysis allows easy visualization of relationships between the entities occupying the various nodes and the intervening edges (**Figure 1**). For this analysis the transplant event, clinical states (acute and chronic GVHD, relapse) and outcomes (TRM, GRFS, RFS and OS) were modeled as vertices, and the edges connecting them were considered to have been weighted by the variables examined in the univariate and multivariate analyses. Since these were calculated for each transitional state and final outcome in the total cohort, these measures are depicted at the nodes/vertices, but in truth their magnitude influences the probability of an individual taking a path along a particular edge. The graph methodology provides an easily visualizable means of assessing the relative effect of different variables in clinical outcomes in HCT where multiple inter-related clinical outcomes need to be examined, potentially simplifying complex data inetrpretation.

**Figure 1.**
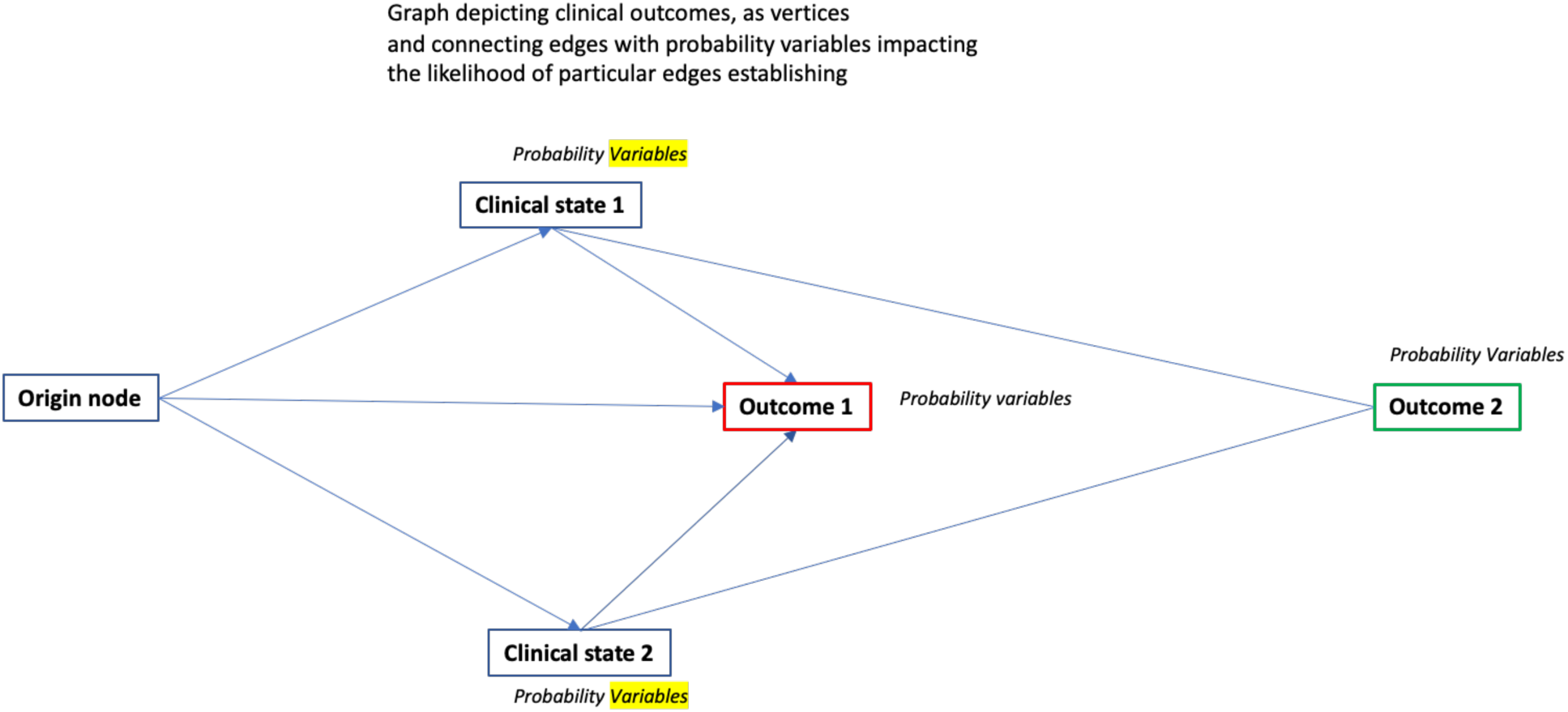
Graph depicting original event, followed by clinical states and outcomes, represented by nodes (vertices) and transition between these nodes represented by edges. The clinical state nodes are first tier transitional states, which lead to the eventual clinical outcome represented by second tier nodes. Edges can be unidirectional or bidirectional, and path along the graph will be determined by prognostic variables. Other intermediary steps may be added as more information becomes available.

## Results

### Patient Demographics

After excluding patients who failed to engraft neutrophils (N = 18) the study cohort was comprised of 686 pediatric donor-recipient pairs (DRP) who underwent unrelated donor HCT for AML (N=355) and ALL (N=333) (**Table 1**). This cohort included pediatric patients aged 0-18 years (average 11 years). All patients received T cell replete grafts; 26% (n = 179) received in vivo T cell depletion, with either anti-thymocyte globulin (ATG) or Alemtuzumab. The majority, 77% received a bone marrow graft and 90% received myeloablative conditioning. The mean(±SD) iKIR, mKIR) and aKIR scores for the entire cohort were 2.5(±1.0), 1.8(±0.85) and 0.5(±0.9) respectively (**Supplementary Figure 1A**). IM KIR score was 5 in 38% donors. All haplotype A donors had an IM KIR score of 4 (**Supplementary Figure 1B**).

### Comparative influence of variables on HCT clinical outcomes, Graph analysis

To put the KIR genotype associated benefit findings in context, graphs depicting clinical outcomes were constructed, consisting of an origin node, constituting the transplantation event. This node had three edges leading to transitory states represented by first order nodes (vertices), accounting for acute and chronic GVHD, as well as relapse. Two additional edges from the origin led to second orders nodes depicting clinical outcomes, TRM and GRFS, in all representing five unidirectional edges emanating from the origin node for this data set. Acute and chronic GVHD also had unidirectional edges leading to TRM and RFS, and a bidirectional edge between the two states. Relapse similarly led to definitive outcome nodes of survival and mortality. The nodes/vertices and edges allowed the clinical events to be clustered in a series of related events (**Figure 2A** **and** **2B**) creating divergent pathways to ultimate survival or demise. Multivariate HR for various demographic and procedural variables were then studied as features of these pathways.

**Figure 2.**
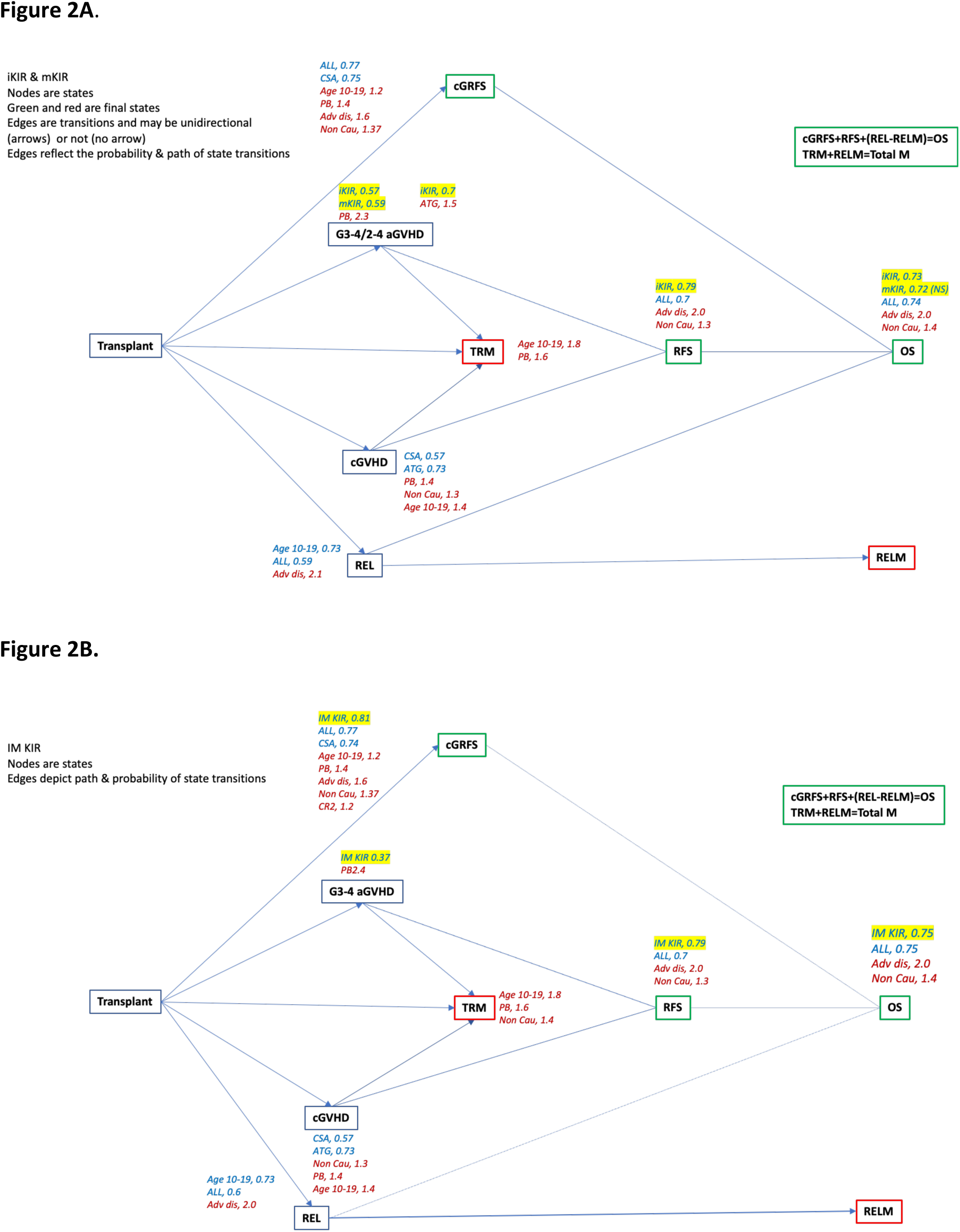
Graph depicting clinical pathways following HCT for pediatric AML and ALL. First tier clinical outcomes include GVHD and relapse, transient states which lead to clinical outcomes, survival, and mortality depicted in second tier nodes. Inhibitory and missing KIR scores (A) and IM KIR scores (B) are depicted.

Variables emerging as significant in the multivariate competing hazards analysis were all encountered on the expected pathway. Advanced disease was the key element in increasing the relapse risk, and downstream decline in RFS and OS, especially in AML patients (**Supplementary Figure 2A**), on the other hand ALL had a favorable impact on relapse risk as well as downstream survival, but in the ALL subset, CR2 fared worse than CR1 (**Supplementary figure 2B**). Older age (10-19 years) along with and non-Caucasian ancestry increased GVHD risk and that of TRM and GRFS. Similarly, PB grafts were associated with increased risk of acute and chronic GVHD, as well as TRM, while ATG diminished the risk of chronic GVHD. Cyclosporine increased the odds for GRFS likely due to a diminution of cGVHD risk (**Figure 2A** **and** **2B**). The pattern of risk factors associated with certain paths (alloreactivity and relapse) seen in the total cohort were largely replicated in AML and ALL (**Supplementary Figure 2A and 2B**).

### Inhibitory KIR score’s impact on clinical outcomes

The calculated KIR scores were examined as a part of the multivariate analysis and their pathway contributions determined. The iKIR score was associated with lowering of the risk of severe acute GVHD, with this benefit of a higher iKIR score extending to RFS and OS in the combined cohort of patients (**Figure 2A**). Similarly, IM-KIR5 led to improved outcomes for all these endpoints as well as for GRFS (**Figure 2B**), essentially identifying a critical role for donor KIR genotyping in transplant clinical outcomes involving alloreactivity in pediatric HLA matched unrelated donor HCT.

Effect of all 3 KIR-KIRL component scores (absolute values of iKIR, mKIR and aKIR) on clinical outcomes evaluated as continuous variables, demonstrated a higher iKIR score was associated with improvement in overall survival (OS), SHR 0.73 (multivariate P=0.003) and relapse-free survival (RFS), SHR 0.8 (P=0.019). Acute grade 3-4 aGVHD was lower in patients transplanted with higher iKIR or mKIR scores (iKIR; SHR 0.58 (P=0.013); mKIR SHR 0.6 (P=0.041)); importantly, relapse rates were not significantly different. In this cohort higher aKIR scores, were not significantly associated with any of the clinical outcomes examined. IM-KIR score was dichotomized for analysis into those DRP with an IM KIR score of 5 (IM = 5), and those with an IM-KIR score of either 2, 3 or 4 (IM < 5). Those transplanted with IM=5 donors demonstrated improved OS, SHR 0.75 (P = 0.027), relapse-free survival, SHR 0.79 (P=0.05) and grade 3-4 aGVHD incidence, SHR 0.37 (P= 0.003) in multivariate analyses, confirming univariate analysis findings (**Figure 3**).

**Figure 3.**
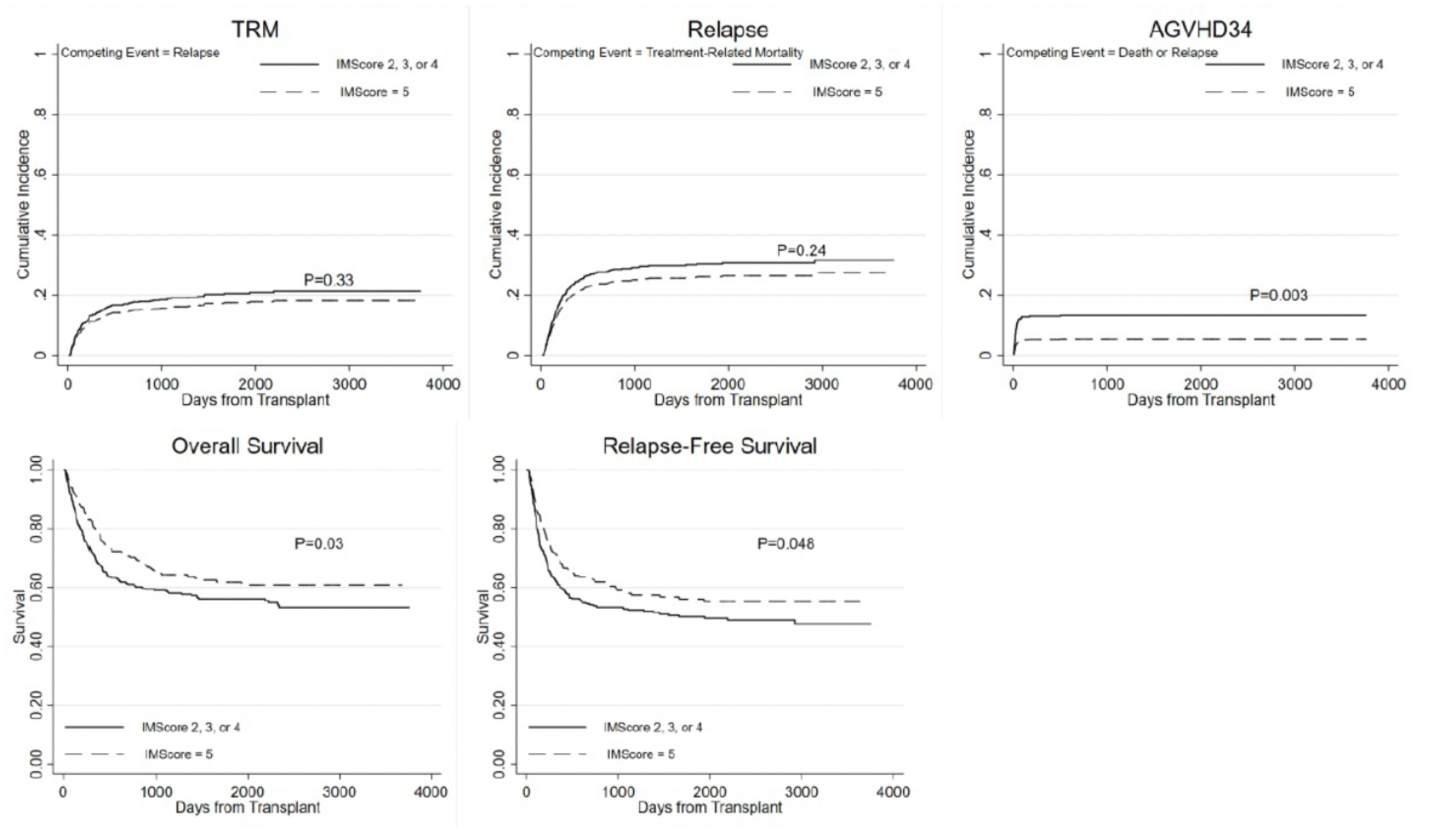
Unadjusted cumulative incidence curves by IM-KIR scores for relapse, treatment-related mortality, and acute grade 3-4 graft versus host disease, as well as failure curves by IM-KIR scores for overall survival, and relapse-free survival. Kaplan-Meier survival curves by dichotomous IM-KIR scores were compared using the log-rank test, cumulative incidence curves were compared using Grey’s test.

### Benefit in AML vs. ALL patients

NK cells are known to most significantly impact clinical outcomes in patients with myeloid malignancies. Therefore, given the GVHD pathway findings of IM-KIR genotype benefit in OS, RFS and grade 3-4 aGVHD in the combined cohort, both the ALL and AML subsets were studied separately to measure any disease-specific effects. Patients transplanted for AML with IM=5 donors saw an improvement in OS, SHR 0.62 (P=0.01), as well as relapse-free survival, SHR 0.70 (p=0.03) in multivariate analysis (**Figure 4**). Relapse was marginally improved in this cohort in univariate analysis SHR 0.67 (P=0.048) however this effect was not verified in multivariate analysis. As noted above, grade 3-4 aGVHD demonstrated a trend toward improvement in outcome, SHR 0.45 (P=0.071) in AML. On the other hand patients transplanted for ALL with IM=5 donors demonstrated a marked decrease in grade 3-4 aGVHD incidence SHR, 0.31 (P=0.017) on multivariate analysis (**Figure 4**).

**Figure 4.**
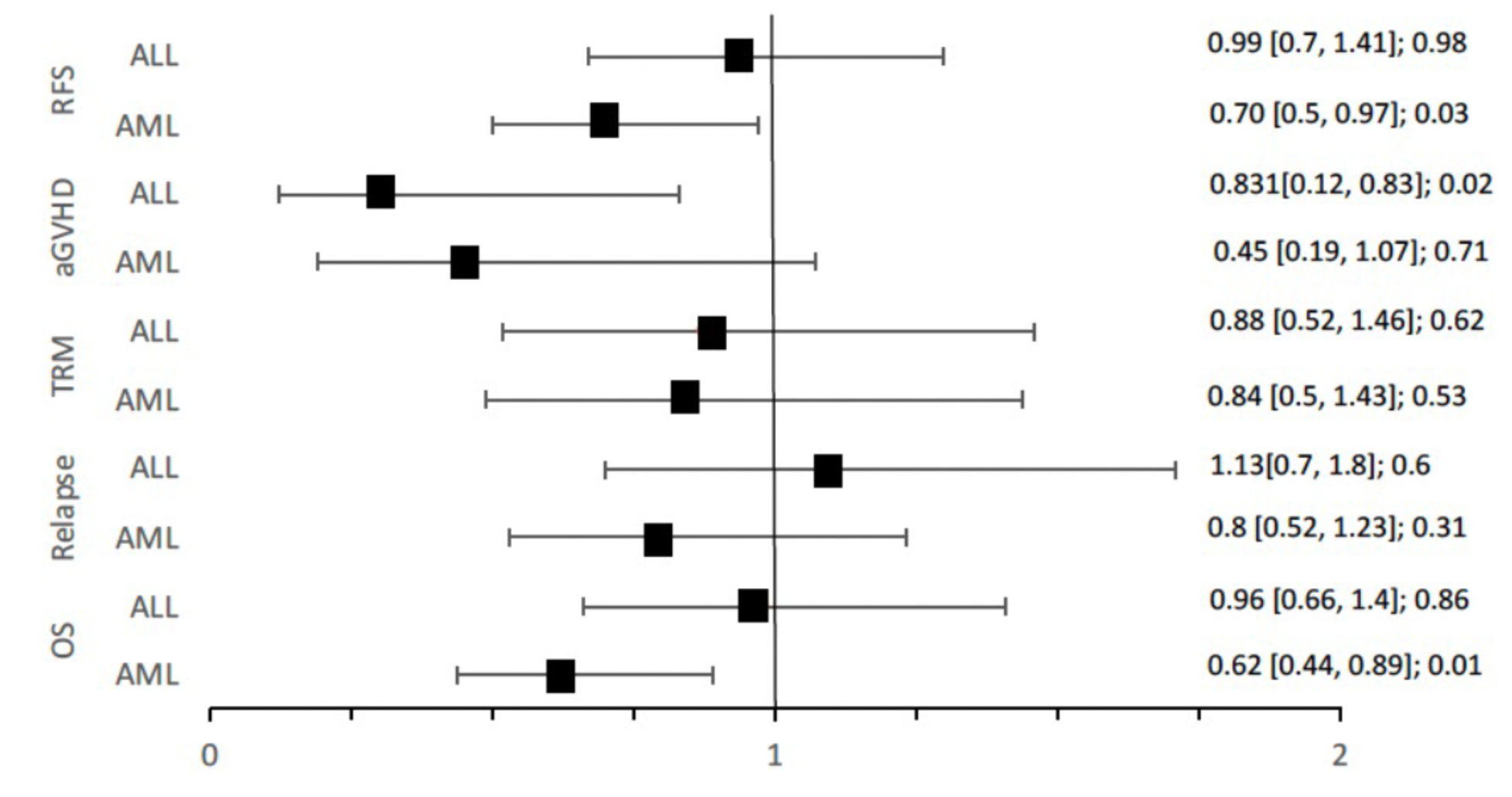
Adjusted sub-distribution hazard and hazard ratios from competing risk and Cox proportional hazard models for the effect of IM=5 scores on clinical outcomes following HCT for AML and ALL.

## Discussion

In this study, the previously published logic-based model examining known KIR-KIRL combinations is validated in a large cohort of pediatric patients who underwent HCT for hematologic malignancy including ALL and AML. In these HLA-matched, unrelated DRPs, those transplanted with KIR haplotype B donors with the highest donor inhibitory KIR content, IM=5, experienced decreased rates of grades III-IV aGVHD and consequently experienced improved RFS, GRFS and OS. These benefits were seen in a patient cohort, where all the standard prognosticators impacted clinical outcomes in an expected direction, such as advanced disease impacting relapse and GRFS likelihood and peripheral blood stem cell grafts demonstrating a greater propensity to develop GVHD with an adverse impact on survival.

Transplant outcomes are complex with multiple complications impacting the clinical course and eventual outcomes. Patients may experience regimen related toxicity, infection, disease relapse and alloreactivity, all of these contributing to eventual survival following the procedure. Traditionally all these clinical endpoints and their risk features are evaluated as discrete entities, measuring the prognostic impact of demographic features on each endpoint. This method limits the ability to develop a deep understanding of the interactions of these prognostic factors as they impact various interrelated clinical outcomes. Graph theory offers a potential solution to this problem, by enabling organization of sequential clinical events as a series of nodes, edges and pathways, to allow study of the cascading effect of different prognosticators for each clinical endpoint (node) and eventually survival. For example, in the pediatric data set examined, the advanced disease was a risk factor for the primary outcome relapse, and downstream RFS, GRFS and OS. Graph-based analysis of post-transplant outcomes may thus yield mechanistic insights into demographic variables studied.

NK cells are considered to exert their protective effect following HCT by enabling graft vs. leukemia (GVL) through NK cell receptors like KIR, CD16 and NKG2A. Inconsistent results have been observed when various KIR molecules are examined individually and in aggregate for their impact on transplant outcomes. When the known KIR-KIRL interactions were summed using a system of vector-operator equations to generate individual donor-recipient scores for analysis, counterintuitively, the presence of a larger number of inhibitory KIR molecules was associated with improved survival in an acute leukemia cohort and lower relapse risk in a very large CIBMTR cohort of AML patients of HLA matched unrelated donor transplants. This study reinforces the notion that increasing number of inhibitory KIR in pediatric transplant recipients may have a beneficial effect, with the novel graph-based analysis demonstrating acute GVHD risk reduction and downstream improvement in related clinical outcomes, RFS and OS, as well as GRFS.

If the positive effect of inhibitory KIR on transplant outcomes is the result of NK cell mediated GVL, only a marginal effect on relapse risk reduction was observed in the pediatric AML cohort, less prominent than in a previously studied adult cohort, where the strongest KIR associated relapse protection was observed in patients who received reduced-intensity conditioning regimens, and in those with in vivo T cell depletion and PBSC grafts, with intermediate risk disease.^8,14^ It was postulated that these patients who receive RIC regimens with ATG highlight the clinic effect of NK cell mediated alloreactivity as, one, relapse rates are highest amongst these patients, and two, in vivo T cell depletion may reduce the T cell mediated GVL effect after HCT allowing NK cell alloreactivity to be evident. However, given the apparent differences in pediatric transplants i.e., the majority of pediatric patients received myeloablative transplants (91%) with bone marrow grafts (77%), and without *in vivo* T cell depletion (26%); it is not surprising that relapse protection is not a dominant effect in this pediatric cohort, especially given the different disease prognosis. The conditions described decreases the risk of relapse in general and change the trajectory of immune reconstitution rendering relapse associations less evident in statistical analysis and explain the relatively modest relapse protection observed here.

Nevertheless, the modest relapse protection effect of iKIR observed deserves discussion, NK cells with a larger number of iKIRs that interact with their cognate self-HLA, produce increased interferon gamma and are more cytotoxic when they meet a non-self-target. This effect is proportionate to the number of iKIR interacting with their self-HLA, a process termed ‘licensing’^5,15^. This effect has been proposed to account for NK cell-KIR alloreactivity in the transplant setting with improved OS and RFS previously. This study’s findings hold true to this and indicate its potential in the pediatric acute leukemia setting. Patients in this cohort transplanted with IM=5 donors experience improvement in the likelihood of developing severe aGVHD. Therapeutic transfer of NK cells has provided insight into NK cell effects in aGVHD, with studies failing to observe aGVHD induction after infusion^16,17^. After HCT, NK cells are the first lymphocytes to reconstitute and increased NK cells at day 14, and after HCT have shown to significantly reduce the development of aGVHD and improve OS. Further, these cells produced significantly more IL-10, IFN-gamma and TNF alpha, especially in the presence of iKIR stimulation^18^. HCT with donors who possess higher inhibitory KIR content results in higher CD56+ counts at day 30 post HCT^7^, consistent with greater proliferative potential of these cells, potentially through a ‘licensing’ mechanism. However, KIR expression on CD56 bright NK cells reconstituting in the immediate aftermath of transplant is low, so a different mechanism is necessary to understand the pathophysiology. Aside from NK cell mediated effects on donor-recipient alloreactivity, it is also important to consider that NK cells may not be the only KIR expressing cell type influencing outcomes after HCT. KIR expression on CD8+ cytotoxic T cells has long been recognized.^27, 28^ Inhibitory molecules expressed on T cells have been shown to dampen cytolytic activity the T cells, and engender tolerance.^29, 30, 31^ Recent studies have shown that the presence of inhibitory KIR, enhanced HLA class I–restricted antiviral CD8 T-cell immunity and outcomes in chronic viral infections, indicating that KIR expressing CD8 T cells might influence the outcome of viral illness^19,20^. All together this presents a plausible explanation for the reduction in aGVHD, where increased iKIR+ CD8+ T cells which may dampen alloreactive aGVHD.

It is interesting to note that reduced acute GVHD was best observed amongst patients transplanted for ALL. Given current and historical practices one may propose that the majority of these patients receive total body irradiation which has the potential to increase the risk of aGVHD^21^. Additionally, the relapse rate in those transplanted for ALL was significantly lower than those transplanted for AML (Chi-Square11 p= 0.0008), resulting in fewer competing events for aGVHD to emerge.

This IM-KIR is a simple logic-based model of assessing potential donor NK cell alloreactivity and needs to be prospectively studied for unrelated donor selection, because of the improved OS and RFS observed for both pediatric AML and ALL, and reduced relapse observed in adults^7,8^. However, it is important to note the limitations of this study including that it is a retrospective cohort study using registry data. In addition, important disease characteristics are missing for this cohort including cytogenetic risk data, and minimal residual disease status prior to transplantation. The incorporation of these characteristics into a prospective trial will better allow us to characterize the outlined potential benefit of KIR-based donor selection in the pediatric HLA-matched setting.

In conclusion, reduced aGVHD, improved survival, relapse-free survival were observed in this large pediatric cohort transplanted with IM= 5 donors for both AML and ALL. This finding was facilitated by using graphs depicting sequential clinical events, leading to outcomes. For the pediatric patients, this influence of KIR was observed despite the lower relapse and mortality event rates observed in pediatric-age patients and the use of myeloablative preparatory regimens with T cell replete bone marrow transplants. Our findings challenge the notion that KIR are irrelevant to pediatric donor selection in both AML and ALL. Future clinical trials evaluating donor selection for MUD HCT should include this measure to determine its value prospectively in a pediatric cohort.

## Data Availability

All data produced in the present study are available upon reasonable request to the authors

## Acknowledgements

The authors gratefully acknowledge Stephen Spellman, MS for help with data organization and for critical review of the manuscript. Institutional Review Board at the Virginia Commonwealth University deemed that since data were completely deidentified, this analysis did not constitute human subjects research and according to Section 45 CFR 46.102(l) of the HHS Regulations for the Protection of Human Subjects did not require IRB scrutiny.

## Authorship

EK: Designed study, developed KIR-KIRL scoring algorithm, analyzed data, wrote paper. RQ: Designed study, developed KIR-KIRL scoring algorithm, performed statistical analysis, wrote paper. AT: Designed study, developed KIR-KIRL scoring algorithm, graph-based analysis method, analyzed data, wrote paper.

---

**Supplementary Figure 1.**
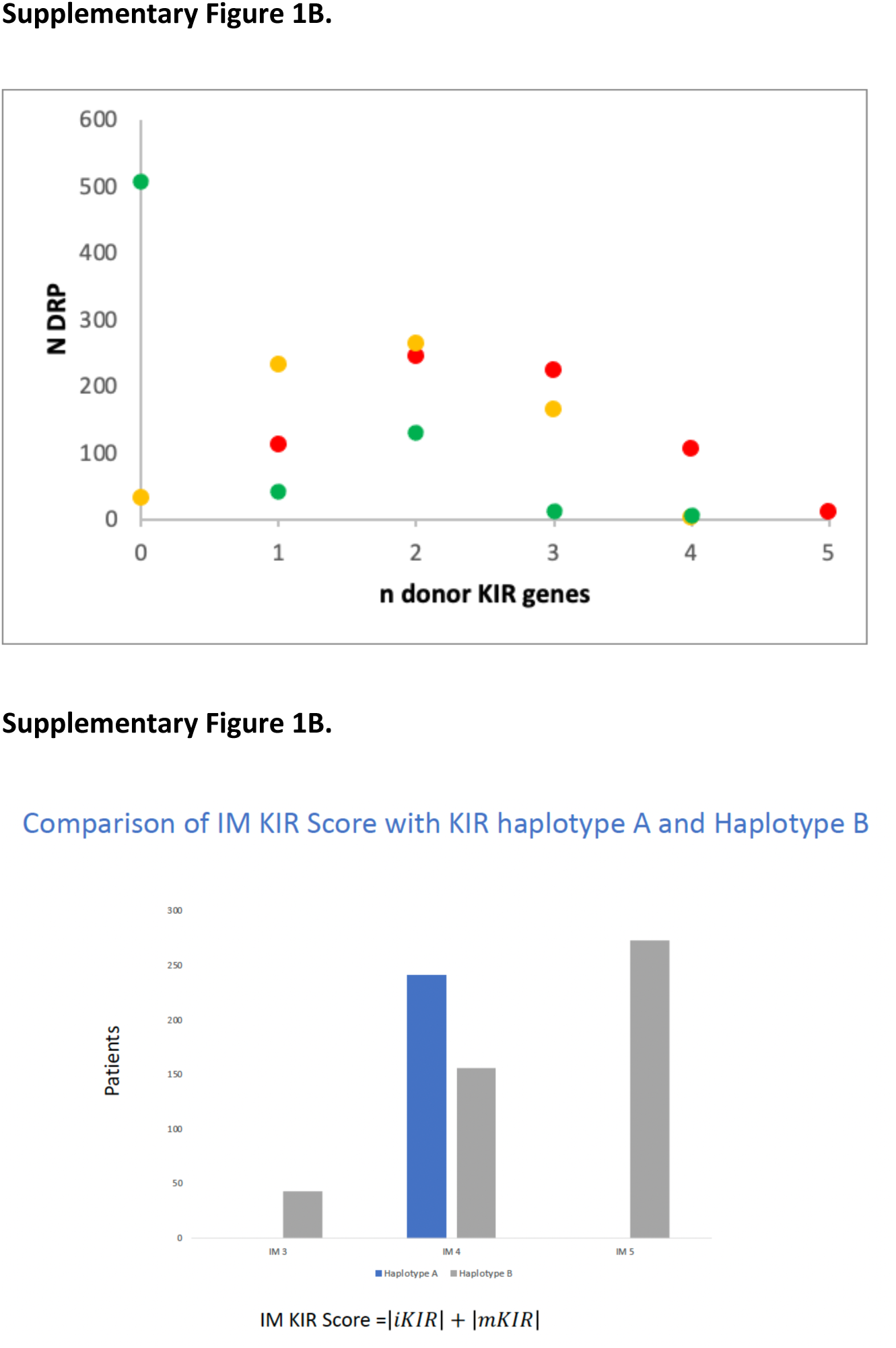
**(A)**Frequency of iKIR (red), mKIR (orange) and aKIR (green) scores, scores on the x-axis and number of donor-recipient pairs (DRP) on the y-axis. **(B)**. IM-KIR score of haplotype A/A and Haplotype B/x

**Supplementary Figure 2.**
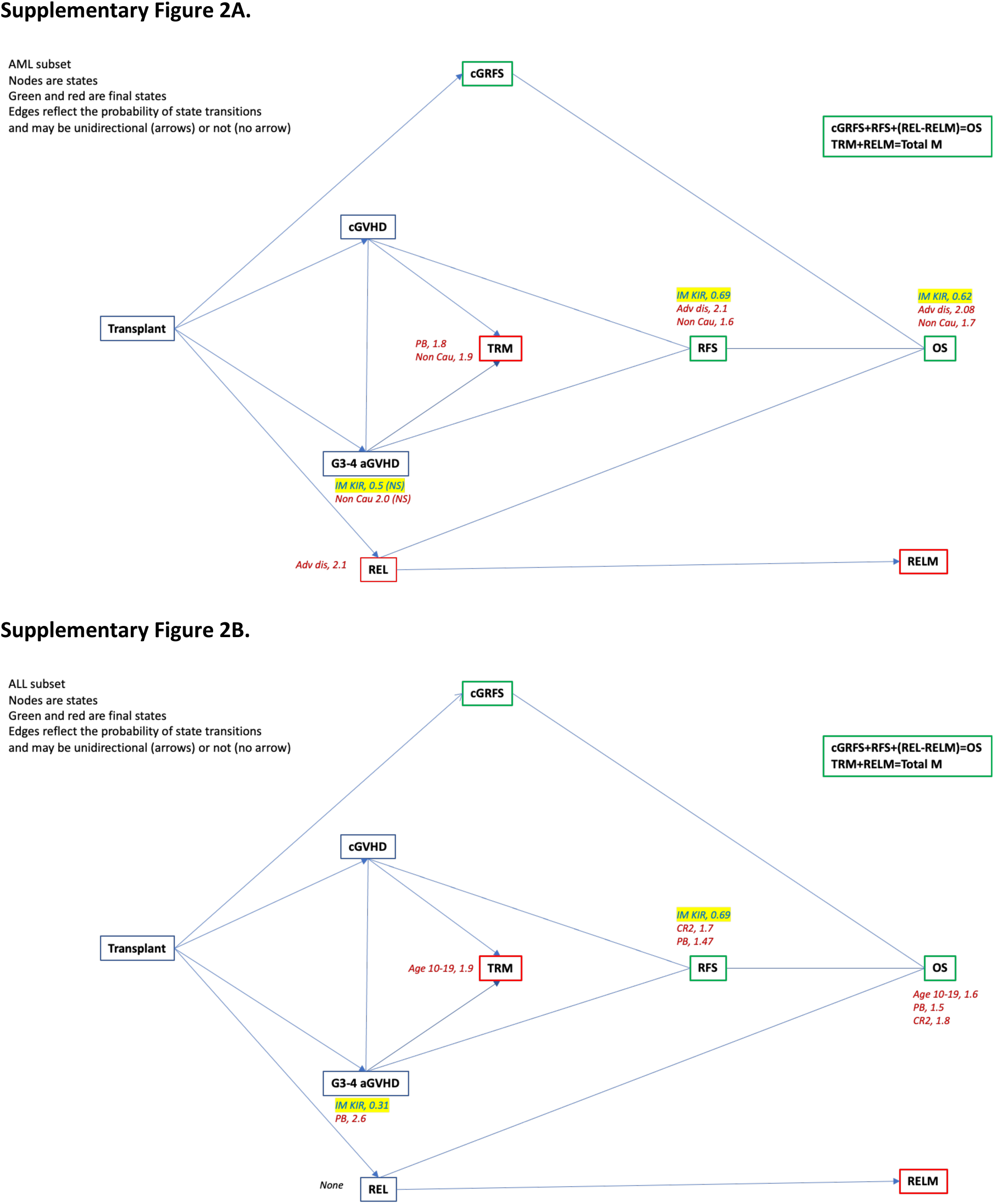
Graph depicting clinical pathways following HCT for pediatric AML (A) and ALL (B). First tier clinical outcomes include GVHD and relapse, transient states which lead to clinical outcomes, survival, and mortality.

